# Genetic diversity among SARS-CoV2 strains in South America may impact performance of Molecular detection

**DOI:** 10.1101/2020.06.18.20134759

**Authors:** Juan David Ramírez, Marina Muñoz, Carolina Hernández, Carolina Florez, Sergio Gomez, Angelica Rico, Lisseth Pardo, Esther C. Barrios, Alberto Paniz-Mondolfi

## Abstract

Since its emergence in Wuhan (China) on December 2019 the Severe Acute Respiratory Syndrome Coronavirus 2 (SARS-CoV-2) has rapidly spread worldwide. After its arrival in South America in February 2020 the virus has expanded throughout the region infecting over 900,000 individuals with approximately 41,000 reported deaths to date. In response to the rapidly growing number of cases, a number of different primer-probe sets have been developed. However, despite being highly specific most of these primer-probe sets are known to exhibit variable sensitivity.

Currently, there are more than 700 SARS-CoV2 whole genome sequences deposited in databases from Brazil, Chile, Ecuador, Colombia, Uruguay, Peru and Argentina. To test how regional viral diversity may impact oligo binding sites and affect test performance, we reviewed all available primer-probe sets targeting the E, N and RdRp genes against available South American SARS-CoV-2 genomes checking for nucleotide variations in annealing sites. Results from this *in silico* analysis showed no nucleotide variations on the E-gene target region, in contrast to the N and RdRp genes which showed massive nucleotide variations within oligo binding sites. In lines with previous data, our results suggest that E-gene stands as the most conserved and reliable target when considering single-gene target testing for molecular diagnosis of SARS-CoV-2 in South America.

## 1. Introduction

The Coronaviridae comprises a large family of pathogenic viruses that are generally transmitted person-to-person through respiratory secretions or fecal-oral route, but may also spread through zoonotic transmission [1]. Members of this family are generally spherical and slightly pleomorphic, ranging from 80-to-120 nm in diameter and covered by distinct projections known as peplomers [2]. Coronaviruses are enveloped positive-sense, single-stranded (ssRNA+) viruses [2] which are known to harbor the largest genomes amongst all known RNA viruses (27-32 kb) [2]. Overall the genomic makeup of COVs embraces a variable number of small open reading frames (ORFs) intercalated among different structural genes (ORF1ab, spike, envelope, membrane and nucleocapsid) defining various lineages [2,3]. Current classification by the Coronaviridae Study Group (CSG) of the International Committee on Taxonomy of Viruses (ICTVs’) recognizes 39 species in 27 genera within the *Coronaviridae* family in the *Nidovirales* order which are known to infect a variety of vertebrates including humans [4,5].

To date, seven coronavirus are known to infect humans. HKU1, NL63, OC43 and 229E which have been associated to mild respiratory and gastrointestinal symptom’s, as well as SARS-CoV, MERS-CoV and the novel emerging SARS-CoV-2 which share similar zoonotic features and are linked to severe disease outcomes often in context of epidemic and pandemic settings [6]. The later three, aside from sharing their host switching capacity and zoonotic origin have also shown a propensity for increased pathogenicity. Such features are intimately related to complex evolutionary mechanisms, including high mutation rates and recombination signals that favor their adaptation process to different species **[2,3]**. After all diversity as seen in most emerging coronaviruses is a feature of host-switching viruses throughout their evolutionary paths. In the last two decades, the spillover of SARS-Coronavirus (SARS-CoV) in China (2002-2003), and MERS-Coronavirus (MERS-CoV) in the Middle East (2012-2016) have shown the deleterious effect of coronaviruses crossing the species barrier, their devastating effects on human health and global impact following pandemic spreads **[7,8]**.

Today, almost two decades after the emergence of SARS-CoV, the newly emerging Severe Acute Respiratory Syndrome Coronavirus 2 (SARS-CoV-2) is now spreading in pandemic proportions after the first reported case in the city of Wuhan, China on December 2019 [9]. This coronavirus-associated acute respiratory disease -Coronavirus 2019 disease- (COVID-19) has turned into a serious global health crisis with 6,407,129 infected individuals and 378,270 reported deaths by June 2^nd^, 2020. As the virus vanishes from most industrialized countries in the northern hemisphere, a new spread wave is sweeping South America under more complex and heterogeneous epidemiological contexts that may favor increased transmission and long-term persistence [10]. In this sense, monitoring SARS-CoV-2 and increased testing capacity becomes a critical issue for most South American nations.

The genome repertoire of SARS-CoV-2 is extensive (<30,000 nucleotides) as most of the Coronaviruses that frequently recombine[11,12]. Its genome alignment is divided into two segment ORF, encoding both non-structural and four structural proteins arranged in the following order spike (S), envelope (E), membrane (M), and nucleocapsid (N). Molecular detection of SARS-CoV-2 relies mainly on three regions with highly conserved sequences: (1) the *RdRp* gene (RNA-dependent RNA polymerase gene) in the open reading frame ORF1ab region, (2) the *E* gene (envelope protein gene), and (3) the *N* gene (nucleocapsid protein gene), with both the *RdRP* and *E* genes exhibiting high analytical sensitivity for detection (technical limit of detection of 3.6 and 3.9 copies per reaction from in vitro transcribed RNA) [13,14], in contrast to the *N* gene which exhibits a much lower analytical sensitivity (8.3 copies per reaction) [14].

While thousands of whole-genome sequences are now available and easily accessible for close to real time visualization in platforms such as NexStrain on the GISAID database (GISAID; https://www.epicov.org) (Elbe and Buckland-Merrett, 2017; Shu and McCauley, 2017), there is a significant shortage on genomic information from SA ever since the virus first arrived in Brazil in March 2019. To date, only 747 genomes from Brazil, Chile, Ecuador, Colombia, Uruguay, Peru and Argentina have been deposited [15]. However, there are no studies addressing genomic diversity and how this may impact molecular diagnosis of SARS-CoV-2. Thus, herein we compared the capacity of all currently accessible primer-probe sets to detect potential SARS-CoV-2 variants by analyzing against all available South American genomes while also assessing regional variation and how this may perhaps predict test performance by screening for potential mutations affecting regions harboring primer-probe binding sites.

## 2. Materials and Methods

The conservation of genomic targets of SARS-CoV-2 for molecular tests was evaluated in this study. As first step a revision about different schemes to detect viral infection by reverse real time PCR was conducted, where we identified sets of primers and probes directed to the E, N and RdRp genes (Table 1). In parallel, whole genome sequences of SARS-CoV-2 reported from South America countries were downloaded from the EpiCoVTM database of Global Initiative on Sharing All Influenza Data (GISAID) [16], the most complete repository of genomic data of pandemic coronavirus causing COVID-19. The complete set of sequences was aligned using MAFFT v7.407 with FFT-NS-2 algorithm and default parameter settings [17,18]. The sequence of Severe acute respiratory syndrome coronavirus 2 isolate Wuhan-Hu-1 was included as reference (Access number NC_045512.2). The multiple alignment obtained from whole genome sequences was used to extract subalignments for ach molecular target (N, E and RdRp genes) in Unipro UGENE v.33.0 [19], considering the annotation of isolate Wuhan-Hu-1 available in National Center for Biotechnology Information (https://www.ncbi.nlm.nih.gov/nuccore/?term=Severe+acute+respiratory+syndrome+coronavirus+2+AND+Whuan). Each sub-alignment was manually verified to remove sequences with Ns, not identified positions (denoted with IUPAC code) and gaps. The “good-sequence” alignment by gene (N, E and RdRp) was used to identify nucleotide and haplotype diversity using DnaSP 5.10 software [20]. A representative sequence by haplotype was used to construct a haplotype alignment and then trees were generated using FastTree double precision version 2.1.10 [21] and visualized in the interactive tool Tree Of Life V4 (http://itol.embl.de) [22]. The genomic regions targeted for each scheme (primers+probe) were subsequently inspected loading the alignment as metadata.

**Table 1.**
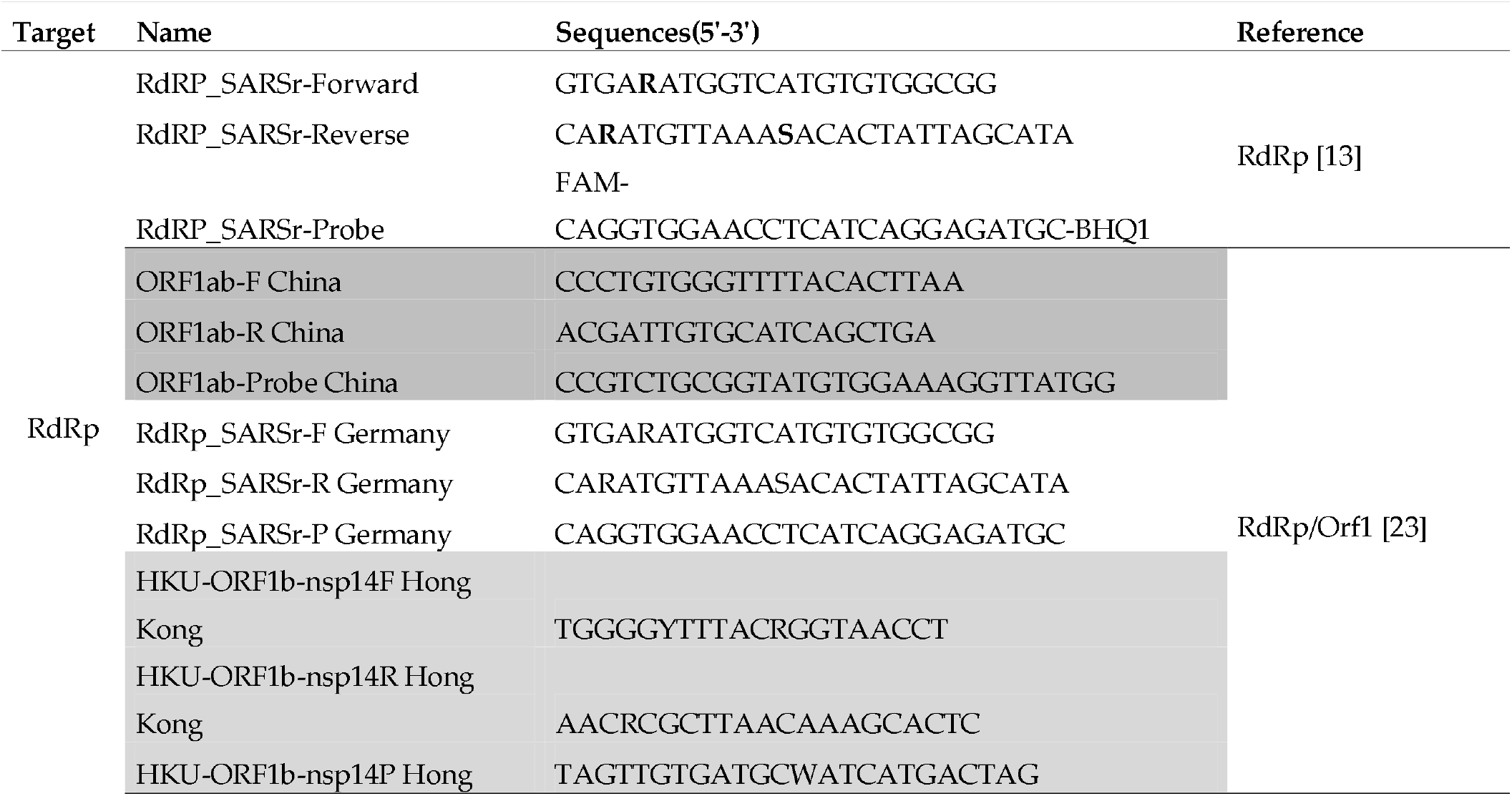
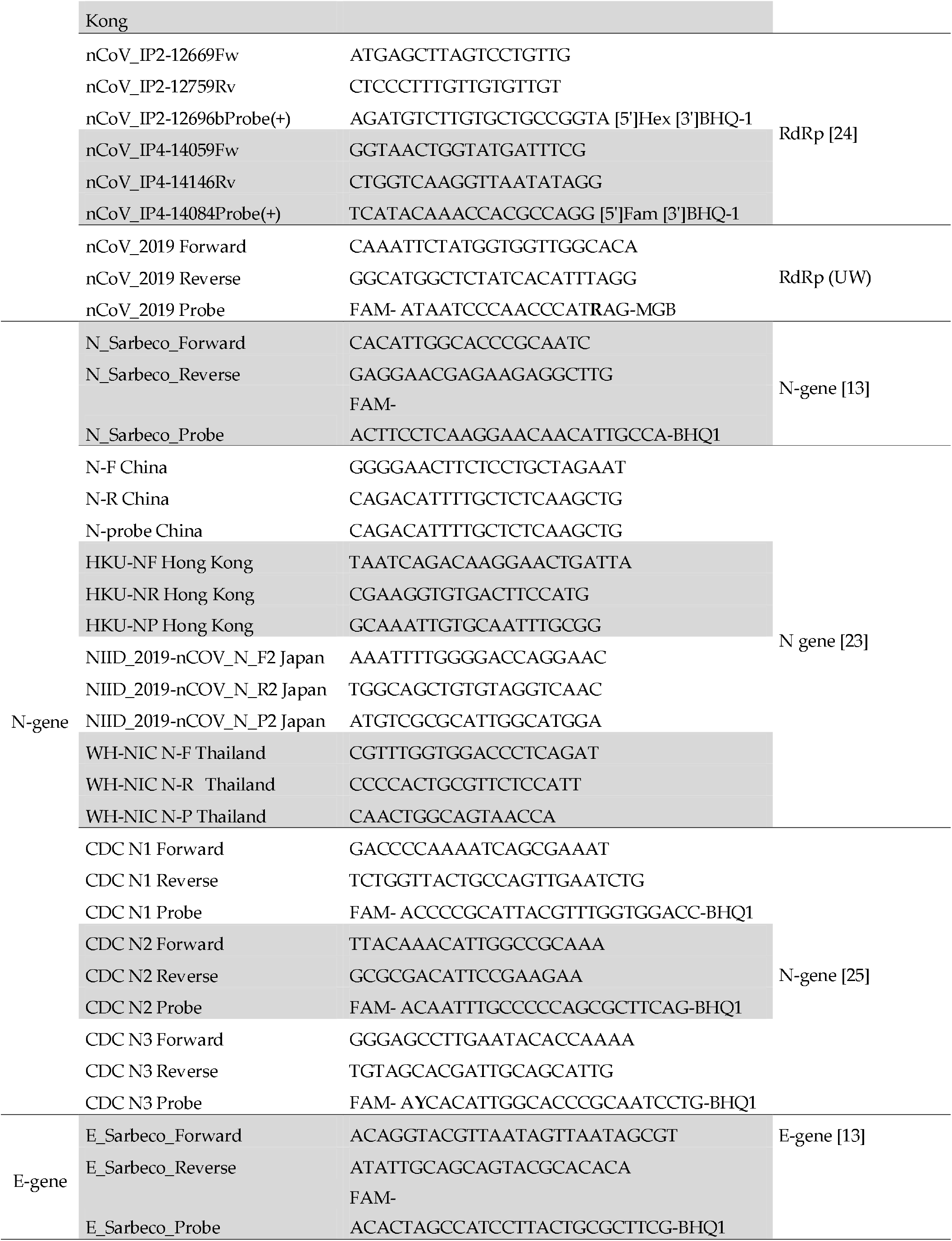
Sequences of primers and probes directed to E, N and RdRp genes and used for the molecular detection of SARS-CoV-2

## 3. Results

A total of 16 oligonucleotide sets (primers+probes) were identified during the revision of schemes to detect SARS-CoV-2 infection using a molecular approach (Table 1). The parallel search of whole genome sequences in GISAID revealed a total of 747, however most of these were duplicated. Verification of redundant sequences showed a total of 373 different genome sequences from the following seven South America countries: Argentina (n= 29), Brazil (n= 95), Colombia (n= 88), Ecuador (n= 4), Peru (n= 2) and Uruguay (n= 11). The results of different diversity parameters evaluated are described in Table 2.

**Table 2.**
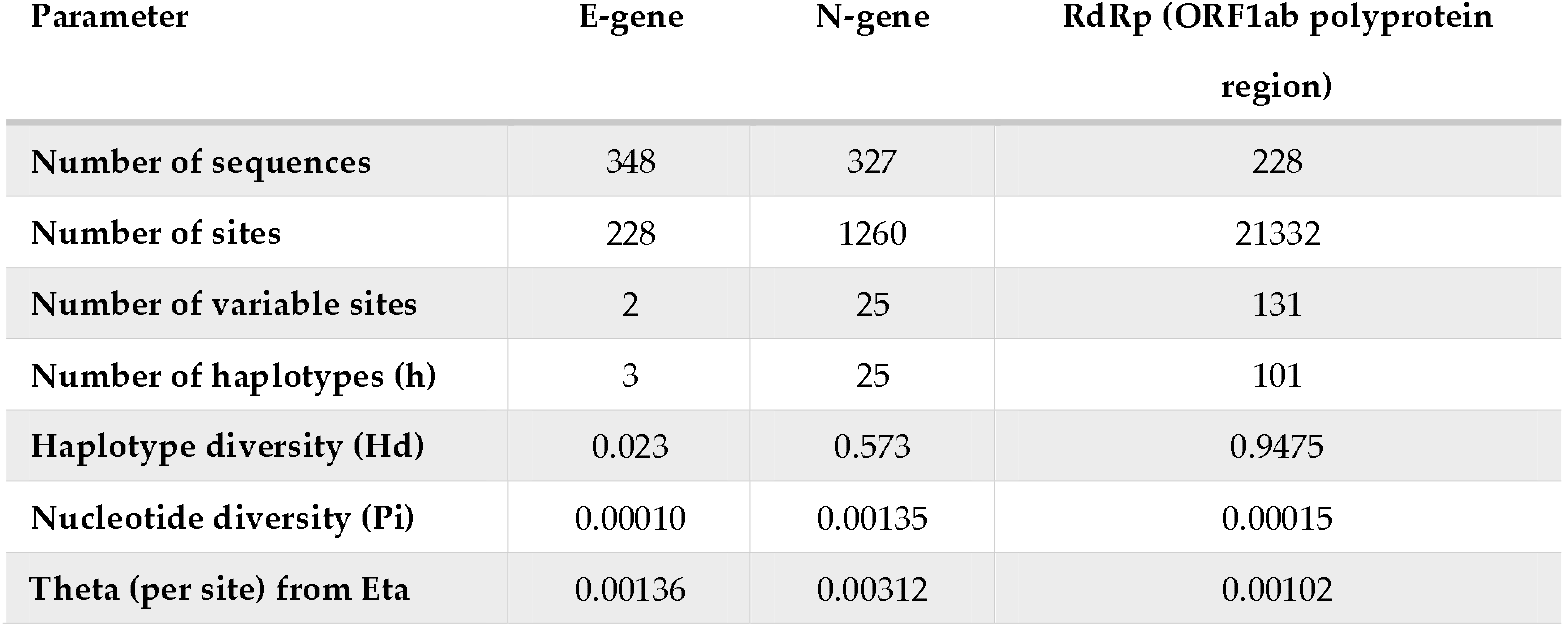
Nucleotide and haplotype diversity of genomic regions of SARS-CoV-2 used for molecular detection

| Parameter | E-gene | N-gene | RdRp (ORF1ab polyprotein region) |
| --- | --- | --- | --- |
| Number of sequences | 348 | 327 | 228 |
| Number of sites | 228 | 1260 | 21332 |
| Number of variable sites | 2 | 25 | 131 |
| Number of haplotypes (h) | 3 | 25 | 101 |
| Haplotype diversity (Hd) | 0.023 | 0.573 | 0.9475 |
| Nucleotide diversity (Pi) | 0.00010 | 0.00135 | 0.00015 |
| Theta (per site) from Eta | 0.00136 | 0.00312 | 0.00102 |

Concerning diversity, in the case of the E-gene, although three haplotypes were identified, only one of them (Hap-1) is predominant in all of the South American countries, with 99.13% of the total sequences (Figure 1A). No variable sites were found in the targets of the scheme flanking this genome region. In the case of the N-gene, a total of 25 haplotypes were identified (Figure 1B), being Hap-2, Hap-1 and Hap-5 the most frequent (with 62.7%, 16.2% and 8.9%, respectively). Variable sites were found in the three oligonucleotides of the CDC-N1 scheme (primers+probe) and in the direct primers of the Jung schemes described in the different countries (China, Hong Kong, Japan and Thailand) (Figure 2). Although most of these variable sites were detected in rare haplotypes, one variable site was detected in a dominant haplotype, involving two nucleotides in the Forward primer of the Jung-China scheme detected in Hap-2. In the case of ORF1ab (RdRp), a total of 101 haplotypes were detected in the analyzed dataset (Figure 1C), being Hap-1, Hap-7 and Hap-30 predominant with 18.1%, 9.3% and 8.8%, respectively. Only in Forward primers of RdRp (Corman) and RdRp/Orf1 (Jung-Germany) were detected variable sites for Hap-64 and Hap-65, and in the reverse primer of nCoV_IP2 (Pasteur) for Hap-5 and Hap-7. All areas with variable sites are marked with a black arrow in Figure 3.

**Figure 1.**
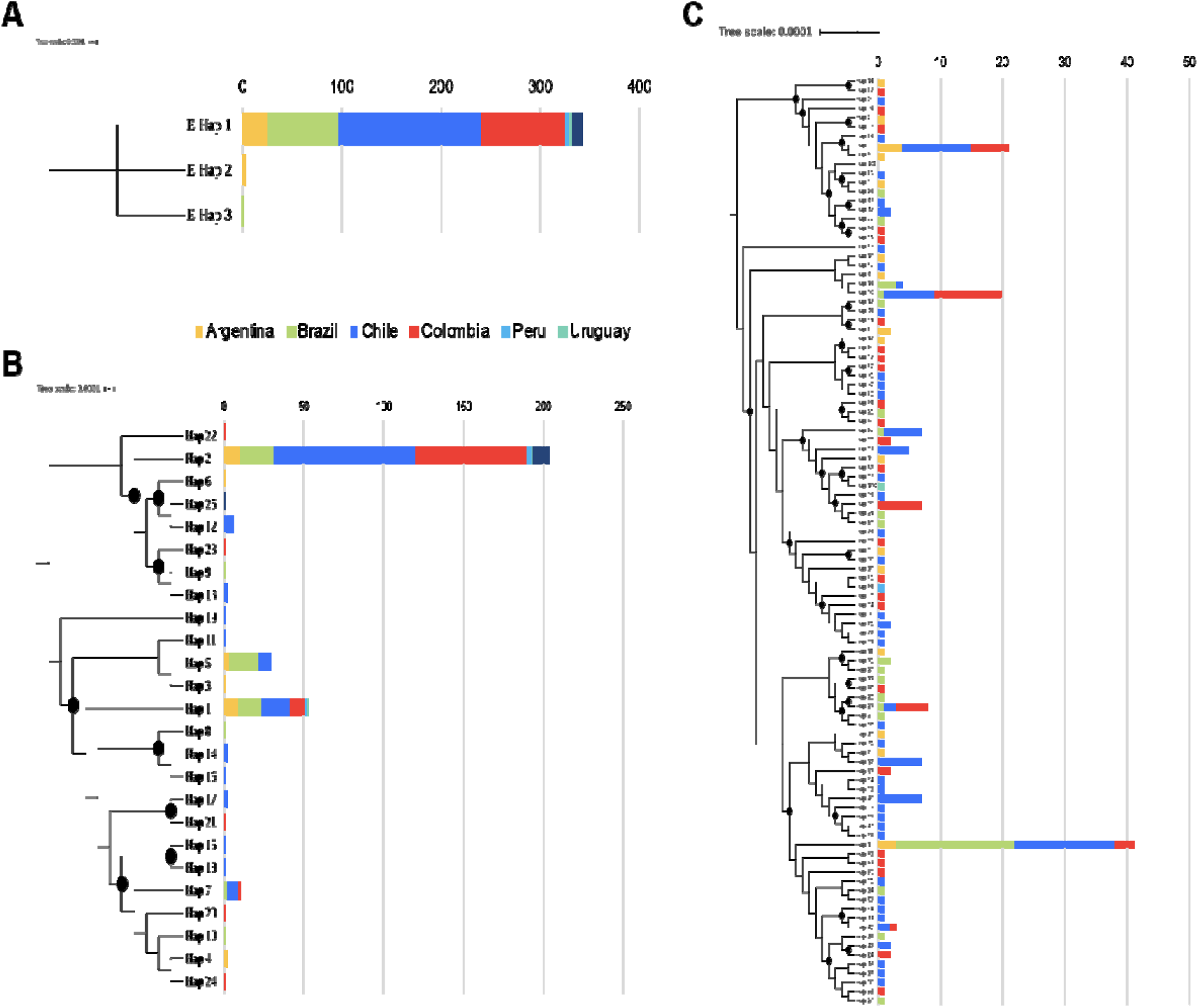
Distribution of haplotypes per gene in the different countries. **A.** Gene E showed only 3 haplotypes and haplotype 1 was the most frequent in the region **B.** Gene N showed 25 different haplotypes, haplotypes 1, 2, 5 and 7 were the most frequent in the region. **C.** Gene RdRp showed 101 haplotypes and the most diverse gene used in the molecular diagnosis.

**Figure 2.**
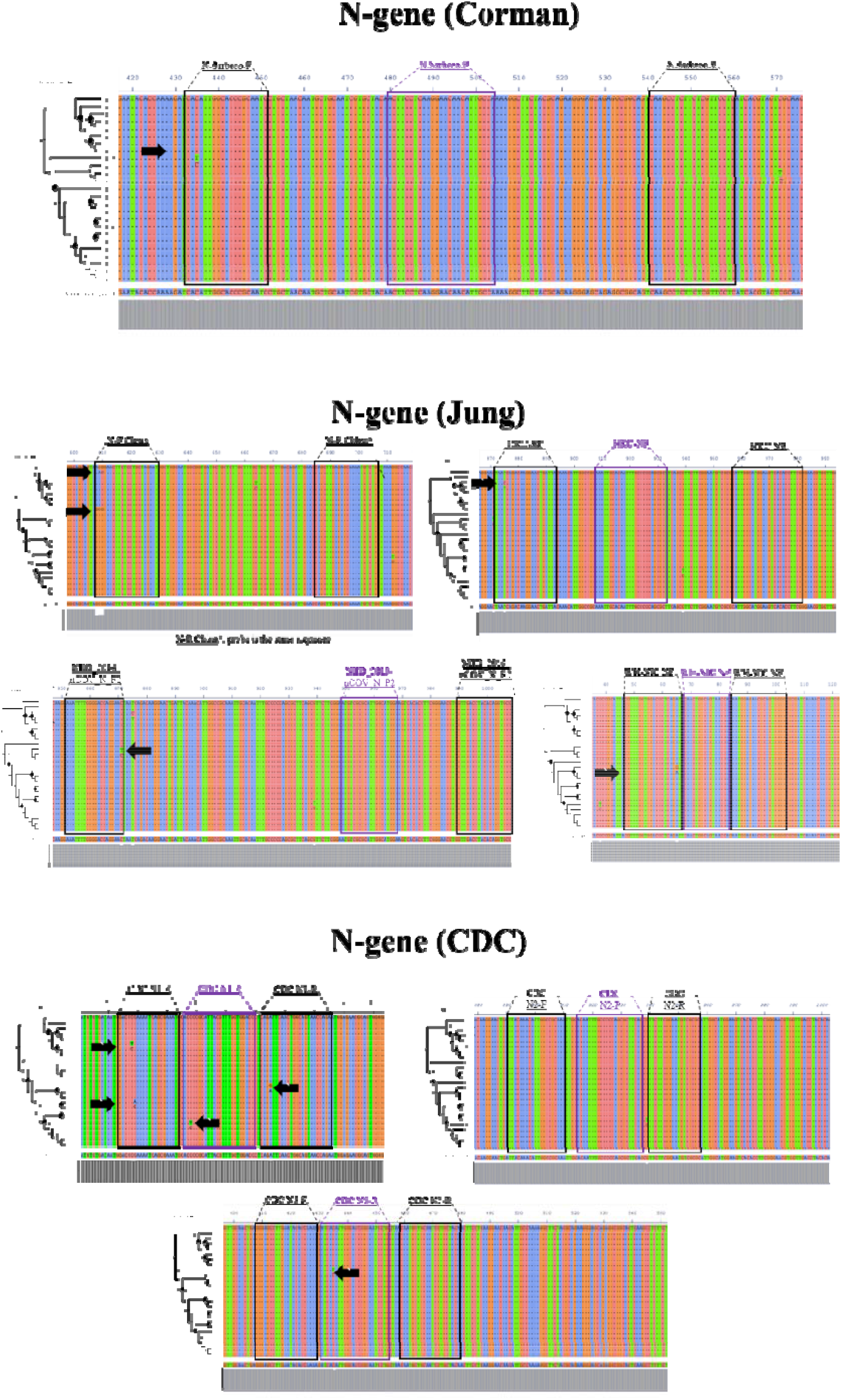
Multiple alignments of the N gene sequences. There were identified several polymorphisms in the annealing regions of the primers of three schemes (black arrows) (Corman, Jung and CDC).

**Figure 3.**
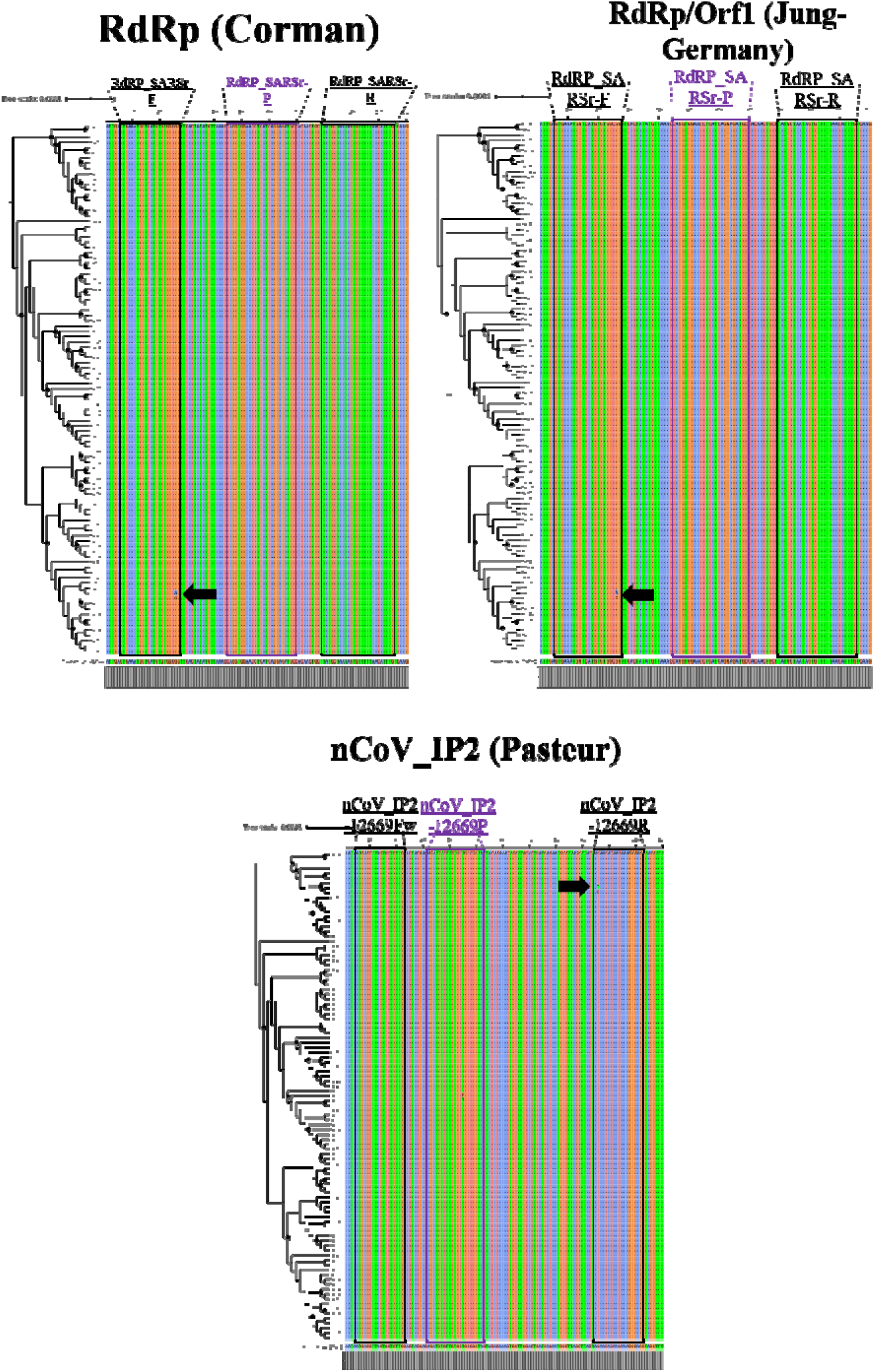
Multiple alignments of the RdRp gene sequences. There were identified several polymorphisms in the annealing regions of the primers of three schemes (black arrows) (Corman, Jung and Pasteur).

## 4. Discussion

SARS-CoV2 has become one of the most important epidemics after the 1917-1918 “Spanish” influenza pandemic [26]with over 7 million infected individuals and close to 400,000 reported deaths worldwide [27]. To date, the majority of affected countries are developed nations with strong public health systems and up-to-date medical facilities, which despite this have undergone severe hassles throughout the course of the pandemic. On the other hand, as the spread wave levels-off in many regions of the world, other regions such as South America are confronting an important rise in case numbers. Many developing countries have been affected by this novel coronavirus, particularly Brazil, Ecuador, Peru and Colombia, which are clear examples were COVID-19 is taking a deadly toll [28].

These countries share many factors in common such as marked poverty, lack of access to basic sanitation services, and inadequate health-care facilities[10]. Such aspects are relevant in understanding the distinct shaping of the course of the epidemics and how it can affect differently transmission dynamics of the virus. For example, the overcrowded environment of slums that make up most of the suburban areas in most South American cities preclude ideal social distancing efforts [10]. In addition, the inability to comply with quarantine measures due to stringent economical reasons, as well as the lack of water and appropriate sanitation policies may favor transmission and halt potential mitigation efforts needed for containment of the virus [10]. This is why alongside strengthening public health capabilities there is an urgent need to better understand and address the main drivers influencing the epidemic spread in such heterogeneous scenario. Measures for assessing these scenarios include evaluating the available diagnostic tools for disease detection in order to improve outbreak response and contention.

In this study, we evaluated whether genomic diversity of SARS-CoV-2 could affect the performance of available primer-probes sets directed to the E, N and RdRp genes, widely used nowadays for the molecular detection of SARS-CoV-2. Our results depict the ample genomic diversity present in the RdRp and N genes which is a shared feature with other members of coronaviridae family (Figure 1; Table 2) [2,3]. In contrast, the E gene, for most of the South American genomes clustered within one haplotype with no SNPs present in these regions where different sets of primers and probes are known to anneal. This finding highlights the great utility of this gene as a screening target for diagnosis of SARS-CoV2 in the region. However, it is important to point out that these primer-probe sets may be susceptible to regional variations of the SARS-CoV2 virus.

A number of RT-PCR tests using different primer/probe sets targeting different regions of the viral genome have been and continue to be developed. The performance of these molecular tests is highly reliant on the primers, probes and reagents used [23]. To date, sensitivities of these nucleic acid amplification tests (NAAT) is variable and in many cases less than optimal [29]. It has been proposed that genetic variations in the SARS-CoV-2 genome may play a role in the observed differences in sensitivity seen amongst different assays.

Current available data, at a global scale, reveals that SARS-CoV-2 has accumulated moderate genetic diversity [30]. As opposed to other RNA viruses, COVs’ exhibit a more modest mutation rate, primarily because of their proofreading capacity and greater replication fidelity [31]. Considering the reported underlying global diversity of SARS-CoV-2 to date, it’s mutation rate has been estimated in 2 of ∼ 6 × 10^4^ nucleotides/genome/year [30] with the majority of these mutations described so far as neutral mutations [32].

Regional variations in SARS-CoV-2 diversity has been recognized as a dynamic process of functional and ongoing adaptation of the virus to the human host [30], as well as a result of multiple and variable sources of introduction to other regions [15]. Such is the case of South America were recent phylogenetic analyses suggest that most viruses have entered from Europe, Oceania and in a less proportion from Asia [15]. Even though sequence variation among SARS-CoV-2 isolates remains moderate, several consistent mutation hot spots have been identified on specific locations that are critical target regions for viral detection.

A comparative analysis study by Jung et al [23] showed that a combination of ORF1 ab (China), N2, N3 (USA) and NIID (Japan) displayed the most sensitive and reliable amalgamation of detection targets[23]. Similarly, Wang et al also reported in the occurrence of mutations in ORF1a (nt8782), ORF8 (nt28144) and N (nt29095) regions, contrasting with the E, 6, 7b regions which exhibited a solid conservation with no mutations detected [23]. Comparable findings by Nalla et al comparing 7 different primer-probe sets also showed that the most sensitive assays where those that targeted the E and N2 genes [33].

Previous studies from South America have also identified changes in the ORF1ab, E and nucleocapsid genes, occurring at the level of specific oligo binding sites which could negatively impact adequate viral identification. Despite the relative conservation of the E-gene [29], as also shown by our study, variants associated with a C-to-T transition at position 26340 have shown to affect testing performance in commercial assays such as the cobas® SARS-CoV-2 (Roche) [34]. The fact that this mutation has emerged independently on various occasions adds a word of caution when relying uniquely on this target region, given the latent possibility of mutations that could potentially limit probe binding and impair amplification [34].

As a matter of fact, the Panamerican Health Organization (PAHO) in their most recent guidelines for the detection and diagnosis of COVID-19 virus infection has stated that although a dual-target testing approach using different genetic markers (E, N or RdRp genes) is recommended, once local circulation is confirmed and widespread though a region, single target testing may be implemented as long as curves and other quality assurance specifications are met appropriately [35]. Amongst all current proposed targets the PAHO recommends the use E or RdRP genes for diagnosis, prioritizing the E gene for single-target testing given its slight higher sensitivity[35]. In light of this recommendation and given the absence of reported SNPs within the E gene from over 300 South American genomes analyzed so far, the E-gene stands like the most promising candidate first-line screening tool for the molecular diagnosis of SARS-CoV2 in the region.

Regarding results for the N and RdRp genes, several haplotypes were identified in the analyzed genomes (Figure 1; Table 2). Of interest, we identified that diversity across all South American SARS-CoV2 sequences apportioned in at least 3 different haplotypes for these two genes, portraying the heterogeneity of the circulating strains throughout the region [16]and the potential role of these proteins in viral pathogenesis [15,36,37]. When screening for SNPs across different primer-probe binding regions, we identified several SNPs capable of altering physicochemical properties of the PCR assays (Figure 2 and 3). This diversity as well as its ability to negatively influence diagnostic test performance is a shared feature with other coronaviruses [29,38]. As mentioned earlier, this is the reason why the occurrence of false negative results in context of ongoing epidemics should be taken cautiously. Evidence from Europe on the occurrence of false negative results for N-gene based assays support our *in-silico* findings [39,40]. Despite no present evidence on false negative results for RdRp gene based assays, its inherent mutation rate (due to environmental pressure and its role as a virulence factor) and having account this gene showed low sensitivity [41] is an aspect that deserves further investigation

It is important to note some limitations in the interpretations of our data. First, because all sequences were retrieved from public databases the accuracy of these sequences could not be entirely verified. In addition, given the most recent emergence of the virus in South America, it is possible that our analysis may represent a snapshot of the most recent evolutionary episodes and does not represent the entire developmental history of the different lineages since introduction. Further studies will be needed to expand our ability to characterize the complete and evolving evolutionary track of the virus.

In summary, this preliminary analysis based on the genomic diversity of SARS-CoV-2 in South America demonstrates how the presence of changes in suggested target regions for primer annealing sites may preclude accurate molecular diagnosis of SARS-CoV-2 when targeting locations within the N-gene region. Our results, confirm the relatively conserved fitness of the E-gene region where no mutations were found, thus making it an ideal candidate for first-line screening in the South America.

Due to the lack of resources and unavailability to acquire reagents and consumables for molecular diagnosis in many areas of South America, the implementation of a single marker assay rises as feasible and cost-effective option for diagnostics in resource depleted countries. Future studies should unveil the diagnostic performance in situ of E gene across multiple geographical regions.

## Data Availability

The data is within the manuscript

## Author Contributions

“Conceptualization, JDR and MM.; methodology, JDR and MM.; validation, CF., SG., LP, CH, EB and AR.; formal analysis, MM, APM and CH.; data curation, JDR and MM.; writing—original draft preparation, JDR, MM and APM.; writing—review and editing, MM, CH, CF, SG, LP, CH, EB, AR, APM.. All authors have read and agreed to the published version of the manuscript.

## Acknowledgments

We thank the Dirección de Investigación e Innovación from Universidad del Rosario for supporting the open access of this manuscript.

## References

1. Cui, J.; Li, F.; Shi, Z.-L. Origin and evolution of pathogenic coronaviruses. Nat Rev Microbiol 2019, 17, 181–192, doi:10.1038/s41579-018-0118-9.

2. Woo, P.C.Y.; Huang, Y.; Lau, S.K.P.; Yuen, K.-Y. Coronavirus Genomics and Bioinformatics Analysis. Viruses 2010, 2, 1804–1820, doi:10.3390/v2081803.

3. Luk, H.K.H.; Li, X.; Fung, J.; Lau, S.K.P.; Woo, P.C.Y. Molecular epidemiology, evolution and phylogeny of SARS coronavirus. Infection, Genetics and Evolution 2019, 71, 21–30, doi:10.1016/j.meegid.2019.03.001.

4. de Groot, R. J. Virus taxonomy: classification and nomenclature of viruses: ninth report of the International Committee on Taxonomy of Viruses; King, A.M.Q.; Ed., Academic Press: London□; Waltham, MA, 2012; ISBN 978-0-12-384684-6.

5. Gorbalenya, A.E.; Baker, S.C.; Baric, R.S.; de Groot, R.J.; Drosten, C.; Gulyaeva, A.A. The species Severe acute respiratory syndrome-related coronavirus: classifying 2019-nCoV and naming it SARS-CoV-2. Nat Microbiol 2020, 5, 536–544, doi:10.1038/s41564-020-0695-z.

6. Andersen, K.G.; Rambaut, A.; Lipkin, W.I.; Holmes, E.C.; Garry, R.F. The proximal origin of SARS-CoV-2. Nat Med 2020, 26, 450–452, doi:10.1038/s41591-020-0820-9.

7. Drosten, C.; Günther, S.; Preiser, W.; van der Werf, S.; Brodt, H.-R.; Becker, S.; Rabenau, H.; Panning, M.; Kolesnikova, L.; Fouchier, R.A.M.; et al. Identification of a Novel Coronavirus in Patients with Severe Acute Respiratory Syndrome. N Engl J Med 2003, 348, 1967–1976, doi:10.1056/NEJMoa030747.

8. Zaki, A.M.; van Boheemen, S.; Bestebroer, T.M.; Osterhaus, A.D.M.E.; Fouchier, R.A.M. Isolation of a Novel Coronavirus from a Man with Pneumonia in Saudi Arabia. N Engl J Med 2012, 367, 1814–1820, doi:10.1056/NEJMoa1211721.

9. World Health Organization (WHO) Novel Coronavirus (2019-nCoV). SITUATION REPORT - 1; 2020;

10. Miller, M.J.; Loaiza, J.R.; Takyar, A.; Gilman, R.H. COVID-19 in Latin America: Novel transmission dynamics for a global pandemic? PLoS Negl Trop Dis 2020, 14, e0008265, doi:10.1371/journal.pntd.0008265.

11. Khailany, R.A.; Safdar, M.; Ozaslan, M. Genomic characterization of a novel SARS-CoV-2. Gene Reports 2020, 19, 100682, doi:10.1016/j.genrep.2020.100682.

12. Sawicki, S.G. Coronavirus Genome Replication. In Viral Genome Replication; Raney, K.D.; Gotte, M.; Cameron, C.E.; Eds., Springer US: Boston, MA, 2009; pp. 25–39 ISBN 978-0-387-89425-6.

13. Corman, V.M.; Landt, O.; Kaiser, M.; Molenkamp, R.; Meijer, A.; Chu, D.K.; Bleicker, T.; Brünink, S.; Schneider, J.; Schmidt, M.L.; et al. Detection of 2019 novel coronavirus (2019-nCoV) by real-time RT-PCR. Eurosurveillance 2020, 25, doi:10.2807/1560-7917.ES.2020.25.3.2000045.

14. Udugama, B.; Kadhiresan, P.; Kozlowski, H.N.; Malekjahani, A.; Osborne, M.; Li, V.Y.C.; Chen, H.; Mubareka, S.; Gubbay, J.B.; Chan, W.C.W. Diagnosing COVID-19: The Disease and Tools for Detection. ACS Nano 2020, 14, 3822–3835, doi:10.1021/acsnano.0c02624.

15. Poterico, J.A.; Mestanza, O. Genetic variants and source of introduction of SARS-CoV-2 in South America. J Med Virol 2020, jmv.26001, doi:10.1002/jmv.26001.

16. Shu, Y.; McCauley, J. GISAID: Global initiative on sharing all influenza data – from vision to reality. Euro Surveill. 2017, 22, 30494, doi:10.2807/1560-7917.ES.2017.22.13.30494.

17. Katoh, K.; Standley, D.M. MAFFT Multiple Sequence Alignment Software Version 7: Improvements in Performance and Usability. Molecular Biology and Evolution 2013, 30, 772–780, doi:10.1093/molbev/mst010.

18. Katoh, K. MAFFT: a novel method for rapid multiple sequence alignment based on fast Fourier transform. Nucleic Acids Research 2002, 30, 3059–3066, doi:10.1093/nar/gkf436.

19. Okonechnikov, K.; Golosova, O.; Fursov, M. Unipro UGENE: a unified bioinformatics toolkit. Bioinformatics 2012, 28, 1166–1167, doi:10.1093/bioinformatics/bts091.

20. Librado, P.; Rozas, J. DnaSP v5: a software for comprehensive analysis of DNA polymorphism data. Bioinformatics 2009, 25, 1451–1452, doi:10.1093/bioinformatics/btp187.

21. Price, M.N.; Dehal, P.S.; Arkin, A.P. FastTree: Computing Large Minimum Evolution Trees with Profiles instead of a Distance Matrix. Molecular Biology and Evolution 2009, 26, 1641–1650, doi:10.1093/molbev/msp077.

22. Letunic, I.; Bork, P. Interactive Tree Of Life (iTOL) v4: recent updates and new developments. Nucleic Acids Research 2019, 47, W256–W259, doi:10.1093/nar/gkz239.

23. Jung, Y.J.; Park, G.-S.; Moon, J.H.; Ku, K.; Beak, S.-H.; Kim, S.; Park, E.C.; Park, D.; Lee, J.-H.; Byeon, C.W.; et al. Comparative analysis of primer-probe sets for the laboratory confirmation of SARS-CoV-2; Microbiology, 2020;

24. Institut Pasteur Protocol: Real-time RT-PCR assays for the detection of SARS-CoV-2.

25. CDC Research Use Only 2019-Novel Coronavirus (2019-nCoV) Real-time RT-PCR Primers and Probes.

26. Taubenberger, J.K.; Morens, D.M. 1918 Influenza: the Mother of All Pandemics. Emerg. Infect. Dis. 2006, 12, 15–22, doi:10.3201/eid1209.05-0979.

27. Worldometer COVID-19 CORONAVIRUS PANDEMIC Available online: https://www.worldometers.info/coronavirus/? (accessed on Jun 9, 2020).

28. Burki, T. COVID-19 in Latin America. The Lancet Infectious Diseases 2020, 20, 547–548, doi:10.1016/S1473-3099(20)30303-0.

29. Wang, C.; Liu, Z.; Chen, Z.; Huang, X.; Xu, M.; He, T.; Zhang, Z. The establishment of reference sequence for SARS-CoV-2 and variation analysis. J Med Virol 2020, 92, 667–674, doi:10.1002/jmv.25762.

30. van Dorp, L.; Acman, M.; Richard, D.; Shaw, L.P.; Ford, C.E.; Ormond, L.; Owen, C.J.; Pang, J.; Tan, C.C.S.; Boshier, F.A.T.; et al. Emergence of genomic diversity and recurrent mutations in SARS-CoV-2. Infection, Genetics and Evolution 2020, 83, 104351, doi:10.1016/j.meegid.2020.104351.

31. Denison, M.R.; Graham, R.L.; Donaldson, E.F.; Eckerle, L.D.; Baric, R.S. Coronaviruses: An RNA proofreading machine regulates replication fidelity and diversity. RNA Biology 2011, 8, 270–279, doi:10.4161/rna.8.2.15013.

32. Cagliani, R.; Forni, D.; Clerici, M.; Sironi, M. Computational Inference of Selection Underlying the Evolution of the Novel Coronavirus, Severe Acute Respiratory Syndrome Coronavirus 2. J Virol 2020, 94, e00411-20, /jvi/94/12/JVI.00411-20.atom, doi:10.1128/JVI.00411-20.

33. Nalla, A.K.; Casto, A.M.; Huang, M.-L.W.; Perchetti, G.A.; Sampoleo, R.; Shrestha, L.; Wei, Y.; Zhu, H.; Jerome, K.R.; Greninger, A.L. Comparative Performance of SARS-CoV-2 Detection Assays Using Seven Different Primer-Probe Sets and One Assay Kit. J Clin Microbiol 2020, 58, e00557-20, /jcm/58/6/JCM.00557-20.atom, doi:10.1128/JCM.00557-20.

34. Artesi, M.; Bontems, S.; Gobbels, P.; Franckh, M.; Boreux, R.; Meex, C.; Melin, P.; Hayette, M.-P.; Bours, V.; Durkin, K. May 3, 2020,.

35. Pan American Health Organization, P. Laboratory Guidelines for the Detection and Diagnosis of COVID-19 Virus Infection, 30 March 2020.

36. Astuti, I.; Ysrafil Severe Acute Respiratory Syndrome Coronavirus 2 (SARS-CoV-2): An overview of viral structure and host response. Diabetes & Metabolic Syndrome: Clinical Research & Reviews 2020, 14, 407–412, doi:10.1016/j.dsx.2020.04.020.

37. Gordon, D.E.; Jang, G.M.; Bouhaddou, M.; Xu, J.; Obernier, K.; White, K.M.; O’Meara, M.J.; Rezelj, V.V.; Guo, J.Z.; Swaney, D.L.; et al. A SARS-CoV-2 protein interaction map reveals targets for drug repurposing. Nature 2020, doi:10.1038/s41586-020-2286-9.

38. Drosten, C.; Chiu, L.-L.; Panning, M.; Leong, H.N.; Preiser, W.; Tam, J.S.; Gunther, S.; Kramme, S.; Emmerich, P.; Ng, W.L.; et al. Evaluation of Advanced Reverse Transcription-PCR Assays and an Alternative PCR Target Region for Detection of Severe Acute Respiratory Syndrome-Associated Coronavirus. Journal of Clinical Microbiology 2004, 42, 2043–2047, doi:10.1128/JCM.42.5.2043-2047.2004.

39. zhou, yunying; Pei; F., Wang, L.; Zhao, H.; Li, H.; Ji, M.; Yang, W.; Wang, Q.; Zhao, Q.; Wang, Y. Sensitivity evaluation of 2019 novel coronavirus (SARS-CoV-2) RT-PCR detection kits and strategy to reduce false negative; Infectious Diseases (except HIV/AIDS), 2020;

40. Su, Y.C.; Anderson, D.E.; Young, B.E.; Zhu, F.; Linster, M.; Kalimuddin, S.; Low, J.G.; Yan, Z.; Jayakumar, J.; Sun, L.; et al. Discovery of a 382-nt deletion during the early evolution of SARS-CoV-2; Microbiology, 2020; 41.

41. Vogels, C.B.F.; Brito, A.F.; Wyllie, A.L.; Fauver, J.R.; Ott, I.M.; Kalinich, C.C.; Petrone, M.E.; Casanovas-Massana, A.; Muenker, M.C.; Moore, A.J.; et al. Analytical sensitivity and efficiency comparisons of SARS-COV-2 qRT-PCR primer-probe sets; Infectious Diseases (except HIV/AIDS), 2020;

